# COVID-19 testing in schools: Perspectives of school administrators, teachers, parents, and students in Southern California

**DOI:** 10.1101/2021.10.14.21265000

**Authors:** Jennifer B. Unger, Daniel W. Soto, Ryan Lee, Sohini Deva, Kush Shanker, Neeraj Sood, Howard Hu

**Author notes:** Corresponding Author: Jennifer B. Unger, Ph.D., Professor of Preventive Medicine, University of Southern California Keck School of Medicine, 2001 N. Soto St., SSB 302, Los Angeles, CA 90089.

## Abstract

**Background:** School-based COVID-19 testing is a potential strategy to facilitate the safe reopening of schools that have been closed due to the pandemic. This qualitative study assessed attitudes toward this strategy among four groups of stakeholders: school administrators, teachers, parents, and high school students.

**Methods:** Focus groups and interviews were conducted in Los Angeles from December 2020 to January 2021 when schools were closed due to the high level of COVID transmission in the community.

**Results:** Findings indicated similarities and differences in attitudes toward in-school COVID-19 testing. All groups agreed that frequent in-school COVID-19 testing could increase the actual safety and perceived safety of the school environment. School administrators and teachers expressed pessimism about the financial cost and logistics of implementing a testing program. Parents supported frequent testing but expressed concerns about physical discomfort and stigma for students who test positive. Teachers and parents noted that testing would prevent parents from sending sick children to school. Students were in favor of testing because it would allow them to return to in-person school after a difficult year of online learning.

**Conclusion:** In-school COVID-19 testing could be a useful component of school reopening plans and will be accepted by stakeholders if logistical and financial barriers can be surmounted and stigma from positive results can be minimized.

## INTRODUCTION

The COVID-19 pandemic necessitated the closure of schools worldwide. By mid□April 2020, 192 countries had closed schools, affecting more than 90% of the world’s student population (Krishnaratne et al., 2020). Closing schools can reduce the spread of COVID-19; however the effect of school closures on COVID-19 transmission and its perceived risks for families with school age children is modest (Prem et al., 2020). Closing schools has detrimental psychosocial, societal, and economic consequences for children and their families (Dimitri A. Christakis, 2020; Dibner et al., 2020). Therefore, reopening schools safely is an important priority.

In addition to physical distancing measures, mask mandates, improved cleaning procedures, and keeping students in small groups, COVID-19 testing in schools could be a useful strategy to identify and isolate infected individuals quickly and restore feelings of safety in school (Krishnaratne et al., 2020). Schools serve as the main trusted point of contact for many families and are conveniently located in neighborhoods. In-school COVID-19 testing could be used to identify positive cases quickly and implement a quarantine strategy promptly to mitigate outbreaks, especially among populations with limited access to quality healthcare and COVID-19 testing (Lewis et al., 2021). Although school-based COVID-19 transmission appears to be rare (Dawson et al., 2021; Ladhani et al., 2021), clusters of transmission do occur (Gold et al., 2021). Identifying, curtailing and monitoring these outbreaks is an important public health priority towards reducing COVID-19 transmission rates.

Several organizations have published recommendations for reopening schools safely (CDC, 2021; NASEM, 2020). However, most of these recommendations emphasize physical distancing, masking, and cleaning rather than COVID-19 testing. Many U.S. states have provided free test kits to schools and obtained statewide CLIA waivers to allow COVID-19 tests to be performed at schools, and online toolkits are available to help schools select an appropriate testing strategy (e.g., CDC, 2021; covidtestingtoolkit.org). Despite the potential usefulness of COVID-19 testing as a strategy to reopen schools, anecdotal evidence by our team in late 2020 indicated that many Southern California schools had not included testing in their reopening strategies, possibly because they perceived school-based testing as too expensive or logistically difficult to implement sustainably.

According to Protection Motivation Theory (Rogers & Prentice-Dunn, 1997), people are motivated to take actions to protect their health when they appraise that the threat is serious (e.g., that the disease is severe and that they are vulnerable to it) and when they appraise that their coping response would be effective (e.g., that the proposed solution would be effective in preventing the disease, and that they have the capability to perform the required actions). Guided by Protection Motivation Theory, we conducted a qualitative study to examine the attitudes of school administrators, teachers, parents, and students toward using COVID-19 testing as part of a strategy to reopen schools. We examined perceived threat of returning to school during the pandemic (perceived severity and perceived vulnerability) and the perceived efficacy of COVID-19 testing as a response to this threat (perceived response efficacy and perceived self-efficacy).

## METHODS

This study used a qualitative approach to assess the feasibility and acceptability of incorporating COVID-19 testing along with other established safety measures to reopen schools. We conducted individual interviews of school administrators and focus groups of teachers, parents, and students as a holistic inquiry into the thoughts and opinions of key stakeholders. The Los Angeles County Institutional Review Board approved the study.

### Participants

Participants were recruited from nine school districts across Los Angeles County that had not already announced plans to implement COVID-19 testing as part of their reopening plans. All of the districts were urban, and their student populations were at least 50% Hispanic/Latinx. These school districts consist of a diverse group of public, private, and charter schools that serve high school students. All participants were involved with high schools (administrators of high schools, high school teachers, parents of high school students, and high school students); however, some of the administrators oversaw middle or elementary schools in addition to high schools, and some of the parents had younger children in addition to a high school student. Research staff contacted the superintendents and principals and invited them to participate in a key informant interview or make a referral to another administrator who could participate. Administrators who agreed to participate (N=9) participated in individual interviews via Zoom. Administrators also were asked to send out a recruiting email to their high school teachers, parents, and students. The email contained a link that teachers, parents, and students could click to participate in focus groups. Teachers and parents who clicked on the link were invited to sign an online consent form and register for a focus group. Students who clicked on the link were invited to register for a focus group after their parents signed an online consent form and the student signed an online assent form. Student focus groups were limited to high school students (ages 14-17). We conducted 9 individual interviews of school administrators, 4 focus groups of teachers (total N=23), 5 focus groups of parents (total N=39) and 2 focus groups of students (total N=13).

### Qualitative data collection procedures

Data were collected between December 2020 and January 2021 online via Zoom. Focus groups lasted approximately 45-60 minutes. Participants received a $20 online gift card after the Zoom session. Focus group facilitators received qualitative focus group training from a qualitative methods expert. Informed consent and youth assent were obtained electronically via REDCap. The interviews and focus groups were conducted with a structured interview guide that was tailored to each group (school administrators, teachers, parents, and students). The focus group facilitator informed the participants that some schools were considering the periodic use of COVID-19 rapid antigen tests for screening as part of their reopening plans, and the researchers were interested in their opinions. Sample questions in the interview guide included the following:

- What needs to happen for [you / your child / students] to feel comfortable returning to school?
- How do you feel about COVID-19 testing as part of the plan to reopen schools?
- How often do you think [you / your child / students] should be tested for COVID-19?
- What would make it easier for you to [get tested / get your child tested / implement a testing program] at school?
- What worries you about testing in schools?
- What do you like about testing in schools?
- Do you know people who would be unwilling to participate in school testing? What would those people say?

### Data analysis

The focus groups and key informant interviews were recorded in Zoom, which automatically produces a transcript. The facilitator and notetaker reviewed the transcripts to correct transcription errors. We then analyzed the transcripts qualitatively using a grounded theory approach, in which common themes emerge organically from the data rather than imposing an a priori theoretical framework on the data. Trained coders reviewed the transcripts and identified recurring themes. The team of coders and investigators reviewed and condensed the initial set of themes into a final list of themes that was then used by all coders. Two different staff members coded each statement in the transcripts into one or more of the themes. These themes were then described qualitatively, including representative quotes to illustrate the themes.

## RESULTS

### School administrators

The school administrators’ quotes clustered into four main themes: readiness to reopen, financial barriers to testing, logistical and staffing barriers, and concerns about student discomfort and stigma. Table 1 shows these themes and representative quotes.

**Table 1.** School administrators

| Theme | Quotes |
| --- | --- |
| Readiness to reopen |  |
|  | Our school is ready. Our classrooms are maximized at about 15 to 16 chairs per class. The classrooms are all outfitted with cameras so that they can teach remotely. Half the kids will be in class and the other half will be at home, and then they flip flop for those days. |
|  | They'll do their temperature checks, they'll do their pre-coded questions. They will get a wristband—the colors will change every day. And then they can go back to class. |
|  | If we're bringing kids on campus, they're going to have to go through a process to get onto campus anyhow. Temperature checks, this questionnaire. They're going to have to go through a process to get on campus. To me, adding in the testing just seems like an extra step to add in. |
| Financial barriers to testing |  |
| | The testing itself is expensive. And I'm not quite sure if it's sustainable. Of course, if it's free, then you provide it. But I don't have funds for \$5 per student. |
|  | Unless LA County is paying for all those and handing them to us. But if they're asking the school or the district to pay for it, there is a pretty substantial cost. |
|  | We can figure out logistics. It's how to pay for it. That becomes a challenge for me. |
| Logistical and staffing barriers |  |
|  | From a school principal's viewpoint, the things that come to mind are scheduling and the amount of time. With children, everything will take two times longer than you think it's going to take. |
|  | Wondering what the logistics would be. How long do you have to wait with your kid, or if kids can just get dropped off and be in a waiting zone, so now we have a whole group of kids. What does that take to monitor the waiting zone to keep the kids from interacting with each other? |
|  | Some don't feel like they're trained or it's part of their job. Staff members, we're asking them to give the test. That's not really in their job description. |
| Stigma and fear |  |
|  | Say a kid walks in, he looks fine. He's tested. Fine. He has a good temperature. Good. No problem. And then I call him out in front of 15, 16 kids, in front of a teacher, saying that he's tested positive. The challenge then is, how do I mitigate that with the rest of the 15 kids or even a faculty member going, "Am I exposed?" and how do I continue that class for the next 30 minutes when the only thing they're worried about is I've been exposed and I don't know what's going to happen next. |
|  | If you're like, "Sorry kid, you have to be all alone in this room while your parent comes to pick you up." There's trauma. |

#### Readiness to reopen

School administrators expressed pride in their schools’ elaborate plans to minimize COVID-19 exposure when their schools reopened. Most of these plans involved physical distancing and sanitizing but not COVID-19 testing. Most had not considered the option of implementing a testing program; instead, they focused on safety precautions that were more familiar to them, such as physical distancing, temperature checks, and symptom questionnaires. Because they had already planned their protocols and believed that their protocols would be effective in preventing transmission, most perceived that their schools were ready to reopen safely without COVID-19 testing, and that COVID-19 testing could be an added inconvenience.

#### Financial barriers to testing

When asked about whether they would be interested in incorporating COVID-19 testing into their reopening plans, administrators expressed concern about the monetary cost of testing and whether they would have the resources to sustain a testing program long-term. They believed that COVID-19 test kits were too expensive for their school or district budgets. Administrators were not aware that they could request free test kits from state or county health departments. Even if COVID-19 test kits were free to schools, the implementation of a testing program still would involve personnel costs.

#### Logistical and staffing barriers

Administrators noted the logistical challenges of testing all students, keeping them physically distanced while waiting to be tested and waiting for the test results, and quarantining students who tested positive. A testing program would require personnel to coordinate the testing, physical spaces for testing and quarantining, and extra time. Administrators worried that they lacked enough staff members who were available and willing to administer COVID-19 tests. They noted that conducting COVID-19 tests was not in their staff’s job descriptions and expected staff to be resistant to this new responsibility.

#### Stigma and student discomfort

Administrators expressed concern about how to handle emotional reactions if a student tested positive and had to be quarantined. They worried about the stigma of testing positive, the fear among students who had been in contact with a student who tested positive, and the trauma of being quarantined in a room alone. Despite these concerns, most administrators acknowledged that COVID-19 testing could be an effective strategy to minimize school closures, if the financial and logistical barriers could be overcome.

### Teachers

The teachers’ comments clustered into three themes: support for frequent testing, level of comfort returning to the classroom, and testing to prevent parents from sending sick children to school (Table 2).

**Table 2.** Teachers

|  |  |
| --- | --- |
| Support for frequent testing |  |
|  | I wouldn't mind testing every day. I think it gives you more reassurance that you're not positive. Testing every day, for me, won't be a problem. I would feel more secure, and if I have to be in the classroom, if students are tested every day, I would feel more comfortable. Otherwise, you never know when they get it. So every day is probably more secure. |
|  | When we started talking about reopening, the one thing that sold every teacher on that committee was the idea of daily testing. |
|  | If it's an intrusive one with a stick way up your nose, that'll be uncomfortable for kids. But if it's just a swab, like brushing your teeth, something as simple as that. It's not a big thing. I think it won't be a problem. I think kids will take it easy. And I think they'll be happy to be at school. I think they'll see the positive in that and be more willing to take the test. |
| Level of comfort returning to the classroom |  |
|  | I feel uncomfortable coming in now, especially because of what you hear in the news, all the spikes. |
|  | I definitely don't feel comfortable going back in person. I still live at home, and I have high risk parents. My dad has high blood pressure. My mom is over the age of 60. My older sister has asthma. I love my job and I love my kids, but I love my family more and I don't feel comfortable going in person. |
|  | My level of comfort stems from the fact that I wrote a lot of the cleaning and disinfecting and safety protocols for our school and have monitored the implementation of those protocols, so I am confident in what we've put together. I'm confident in our teachers and our custodial staff to maintain a clean and sanitary environment. I think my comfort with those pieces is because that's what I have direct control over. |
| Testing to prevent parents from sending sick children to school |  |
|  | I have some concerns over things that I don't have control over such as the truthfulness of families or what other people are choosing to do outside of the school environment |
|  | I don't know to what extent I can trust people. You see people in social media gathering, teachers gathering....you would hope that these are the people that are definitely taking care of themselves and they're not. |

#### Support for frequent testing

Teachers were generally in favor of frequent COVID-19 testing as a way to keep the school community healthy and to provide reassurance that the school was safe. They believed that frequent testing would help everyone in the school community feel safer, and that the students could handle frequent testing.

#### Level of comfort returning to the classroom

Many teachers stated that they did not feel safe returning to the classroom. They mentioned that the COVID-19 rate still seemed too high, that there were numerous large gatherings in their community, and that they did not want to infect their own family members. They prioritized the health of their vulnerable family members above the benefits of returning to the classroom. However, some teachers expressed confidence in their school’s ability to implement protocols to protect students and staff from COVID and were proud of their schools’ elaborate preparations.

#### Testing to prevent parents from sending sick children to school

Some teachers believed that testing would make the school environment safer because it would prevent parents from sending their sick children to school. They believed that students and their families were not taking precautions, and that testing would ensure that sick children stayed home. Frequent COVID-19 testing could serve as an added protection against a COVID-19 outbreak originating from families or other staff who were not being careful outside of school.

### Parents

Parents’ opinions clustered into four themes: support for frequent testing, physical discomfort and stigma, testing to prevent parents from sending sick children to school, and concerns about children’s mental health (Table 3).

**Table 3.** Parents

|  |  |
| --- | --- |
| Support for frequent testing |  |
|  | It gives me peace of mind knowing my child would be getting tested and the other children as well. I would allow them to get tested every day, if needed, or twice a day before and after. |
|  | With the caveat that budget is not a concern, then clearly everyday testing is what people would want. |
|  | Testing should be done in the morning. You want to do it on Mondays if you're going to do it only a few times a week. |
| Physical discomfort and stigma |  |
|  | What would be the effect on my daughter? Would she be traumatized? |
|  | Would it be in private? You want to avoid any stigma like someone saying, "Hey, Jimmy tested positive," or something like that. |
|  | When they return back to school, they can get made fun of. I'm just worried that this might bring some sort of negative implication to their self-esteem if they get called out for being positive. |
| Testing to prevent parents from sending sick children to school |  |
|  | Parents still send their kids with a cold because they don't want to miss work or they don't have a babysitter, or whatever the reason. |
|  | I've been in the classroom. I've been in carpools with kids that are coughing and sneezing all over and here we are taking them to basketball practice with my son in the car. But now it's a little bit more serious. |
| Concerns about children's mental health |  |
|  | He's not as engaged as he was before. He says, "I want to go to school. I want to see my friends." He doesn't want to just be on the tablet on zoom. |
|  | They're just not their normal happy. They're still happy kids, really good kids, but I can just tell them this has put a little bit of a strain on them. |
|  | I really wanted her to go back, but she's actually had a lot of anxiety about being surrounded by too many people. Sometimes it's even been a struggle for her to get out of the car so we can go for a walk by the beach. |

#### Support for frequent testing

In general, parents were comfortable with frequent testing in schools. Most were in favor of daily testing, preferably in the morning before the students interacted with one another in the classroom. If daily testing was infeasible, they supported testing several times per week. They reported that frequent testing would bring them peace of mind and make them less worried about sending their children back to school.

#### Physical discomfort and stigma

Parents worried about the physical discomfort of testing and the stigma of testing positive for COVID-19. They worried that their children would get bullied or socially ostracized if they tested positive, and that the testing process could cause physical and emotional trauma.

#### Testing to prevent parents from sending sick children to school

Echoing the concerns of the teachers, parents also expressed worry that other parents would send their sick children to school, and that children would not follow physical distancing guidelines consistently. Some believed that COVID-19 testing would reduce the likelihood that parents would try to bring sick children to school. Parents understood that other parents often send their children to school or other activities when they had minor illnesses, but this was no longer acceptable during a pandemic.

#### Concerns about children’s mental health

Parents also expressed concern about their children’s mental health. Parents reported that their children felt lonely, depressed, and anxious during the pandemic. Some said that returning to school would be beneficial to their children, but others stated that their children had anxiety about returning to in-person learning and preferred to stay remote.

### Students

Students’ comments clustered into three themes: support for frequent testing, concerns about physical discomfort, and opinions about online vs. in-person school (Table 4).

**Table 4.** Students

|  |  |
| --- | --- |
| Support for frequent testing |  |
|  | I would want to get tested every single day, just because it's the most responsible thing to do. I would want them to test every student every day to make sure no positive student gets in. |
|  | Maybe three times a week just due to the fact that testing can be expensive and very difficult and time consuming. |
|  | I'd rather have to be tested every day, than stay at home for another five months. |
| Physical discomfort |  |
|  | Some people say it hurts. Since it would be my first time and I've never taken it before, I would definitely be a little bit nervous. |
|  | Having the ones that go all the way up [the nose], I don't know if that could be a solution for everyday kind of testing. But I am totally okay with any of the swab ones, because I think that's something people can just get used to. |
|  | I think the better one to do would be a throat swab or the spit ones, because I think, because those would be a lot easier for the students to manage comfortably. |
| Online vs. in-person school |  |
|  | I like distance learning because I think it's easier. The teachers do a lot of open book tests which is easy. Distance learning just gives you a bit of independence, to be able to control what you have to do. |
|  | You wake up and sit in front of your computer for six hours. When you actually went to school each day was a little different and more exciting or surprising. |
|  | I don't personally like distance learning just because it's so mentally exhausting having to sit at a desk for eight hours all day. And you don't have any motivation. If we were in physical school, we would have the teachers telling us to do our work and we would have our friends, but now it's just like you become so distant from people, because all you do is sit at a desk all day and do work. |

#### Preferred testing frequency

Overall, students believed that COVID-19 testing in school would be beneficial to prevent COVID-19 transmission and help students feel safe at school. Most students were willing to endure the inconvenience and discomfort of daily testing if it meant that they could go back to school. They viewed testing as a more preferable alternative to staying home.

#### Concerns about physical discomfort

Some students expressed concern about physical discomfort of testing, referring to deep nasal swabs. Most students had either experienced the physical discomfort of a deep nasal swab or had heard about it, and they did not want to have that uncomfortable experience frequently. They preferred less invasive tests such as shallow nasal swabs or saliva tests. As long as the test was not painful, they were willing to be tested frequently.

#### Opinions about online vs. in-person school

Students expressed mixed feelings about returning to in-person learning vs. remaining online. Some students preferred online learning because they perceived it as easier and they liked having more control over their schedules. However, other students expressed frustration with the monotony of online learning and the difficulty of concentrating without a teacher present. Although many students disliked online learning, most agreed that it was a better option than returning to school and potentially exposing their family to COVID. Students expressed sadness about missing their friends and missing out on school activities and milestones.

## DISCUSSION

This qualitative study of attitudes toward in-school COVID-19 testing revealed important similarities and differences in opinions across different types of stakeholders—school administrators, teachers, parents, and students. There was a broad consensus across stakeholder groups that solutions were needed to allow students to return to school safely. All groups agreed that in-school COVID-19 testing could increase both the actual safety of the school and the perception of safety in the school by identifying and isolating infected students quickly, if the logistical barriers could be overcome. There also was consensus among adults about the potential trauma of testing positive, being exposed to a student who tested positive, or quarantining; interestingly, the students themselves were less worried about this trauma than the adults were. The students were willing to assume these burdens to protect one another and their families and to be allowed to return to school.

School administrators and teachers also acknowledged the benefits of COVID-19 testing in general, but they were pessimistic about the feasibility of implementing a testing program in their schools because of financial and logistical barriers. Their fatalism about not being able to implement a testing program seemed to outweigh their desire to explore in-school testing as an option. Indeed, there are numerous barriers to implementing an in-school testing program, including obtaining test kits and CLIA waivers to perform them in a community setting, finding enough staff who are willing and able to perform the tests, allocating rooms and time for testing during the school day, keeping students physically distanced while waiting to be tested and waiting for test results, quarantining students who test positive, and handling emotional reactions of students who test positive and others who had contact with them. It is not surprising that school administrators and teachers viewed these barriers as formidable.

Our study revealed a lack of trust among teachers and parents. Both groups were aware that parents sometimes send sick children to school when they lack childcare. Although this might have been a minor annoyance when sick children transmitted colds and flus to other students, teachers and parents were unwilling to tolerate this level of risk during the COVID-19 pandemic. It might be necessary for schools to insist that students with symptoms stay home. Short-term childcare solutions are needed for sick children whose parents need to work.

Several studies have found that routine testing can reduce COVID-19 transmission in schools (Lanier et al., 2021). Although the prevalence of symptomatic students in schools may be rare, the cases that do occur tend to lead to household transmission (Hershow et al., 2021). If they can overcome the financial and logistical barriers, schools should consider adding frequent COVID-19 testing to their protocols. Although some guidelines and recommendations for school-based testing exist (CDC, 2021; NASEM, 2020), additional research will be needed to identity the optimal testing schedule under varying conditions (e.g., community transmission rate, school size, vaccination status of students and teachers).

This study was conducted in Los Angeles County, California, which has an especially high proportion of Hispanic/Latinx residents compared with the U.S. population overall. In California, Hispanic/Latinx residents are more likely to live in overcrowded households and to be essential workers who must interact with the public (Contreras et al., 2021; Reitsma et al., 2021). This group also lags behind non-Hispanic whites in COVID-19 testing and vaccination (CDC 2021; Reitsma et al., 2021). Locating COVID-19 testing (and perhaps vaccination) services within schools, where families and children go every day, could be an effective strategy to reduce COVID-19 transmission among Hispanic/Latinx families.

### Limitations

This was a convenience sample of school administrators, teachers, parents, and students who responded to an email invitation. Their opinions might not generalize to the overall population. Data were collected in December 2020-January 2021, when the COVID-19 prevalence was peaking, COVID-19 vaccines were not yet available to adolescents, and Los Angeles County schools had not yet reopened. Attitudes toward returning to school with or without COVID-19 testing might have changed since the data were collected. This study focused on high school administrators, teachers, parents, and students. These findings might not generalize to elementary and middle school settings.

### Policy and Practice Implications

Keeping schools open safely is a shared goal of school administrators, teachers, parents, and students. COVID-19 testing has the potential to identify infected students and staff early and prevent them from transmitting the virus in school. Simulation studies have indicated that frequent in-school COVID-19 testing could prevent school-based outbreaks (McGee et al., 2021). However, for schools to implement testing programs, they need to consider numerous factors. They must select which type of test to use (e.g., rapid antigen tests that deliver results within 15 minutes but have lower sensitivity vs. PCR tests that are more sensitive but do not provide quick results) and the feasibility of testing (e.g., availability of test kits, staff to conduct the tests, and rooms to quarantine students who test positive). It is also important to devise testing protocols that protect confidentiality and minimize physical and emotional trauma. This could include teaching students stress management and coping strategies, devising nonthreatening and unobtrusive ways to inform students and parents of positive test results, and providing clear guidance about when students can return to school after a positive test. Schools also must consider how to handle families that do not consent to testing, whether they should adapt the testing schedule according to the weekly or monthly COVID-19 prevalence, how vaccination will affect the need for testing, and whether it is more beneficial to allocate scarce resources to testing or to vaccination. It is likely that some districts and schools will have the resources and training to implement testing programs, and others will need technical assistance, choose to outsource testing to private companies, or choose not to incorporate testing.

## CONCLUSION

This qualitative study of four types of school stakeholders—administrators, teachers, parents, and students—found that most stakeholders prefer in-school COVID-19 testing, and that the main barriers to testing were logistical and financial. If those barriers could be overcome, most of these stakeholders would prefer in-school testing as part of a strategy to reopen schools safely. Efforts are needed to educate school personnel about the resources available to them including testing toolkits, sources of free test kits, and statewide or countywide laboratory waivers and apps to report results. Even with these resources, many schools will still need more personnel and technical assistance to implement testing programs effectively.

## Data Availability

All data produced in the present study are available upon reasonable request to the authors.

## REFERENCES

Centers for Disease Control and Prevention. (n.d.). CDC COVID Data Tracker. Centers for Disease Control and Prevention. https://covid.cdc.gov/covid-data-tracker/#vaccination-demographic.

Centers for Disease Control and Prevention. (n.d.). Operational Strategy for K-12 Schools through Phased Prevention. Centers for Disease Control and Prevention. https://www.cdc.gov/coronavirus/2019-ncov/community/schools-childcare/operation-strategy.html?CDC_AA_refVal=https://%3A%2F%2Fhttp://www.cdc.gov%2Fcoronavirus%2F2019-ncov%2Fcommunity%2Fschools-childcare%2Fschools.html.

Contreras, Z., Ngo, V., Pulido, M., Washburn, F., Meschyan, G., Gluck, F., Kuguru, K., Reporter, R., Curley, C., Civen, R., Terashita, D., Balter, S., & Halai, U.-A. (2021). Industry Sectors Highly Affected by Worksite Outbreaks of Coronavirus Disease, Los Angeles County, California, USA, March 19–September 30, 2020. Emerging Infectious Diseases, 27(7), 1769–1775. https://doi.org/10.3201/eid2707.210425

Dawson, P., Worrell, M. C., Malone, S., Tinker, S. C., Fritz, S., Maricque, B., Junaidi, S., Purnell, G., Lai, A. M., Neidich, J. A., Lee, J. S., Orscheln, R. C., Charney, R., Rebmann, T., Mooney, J., Yoon, N., Petit, M., Schmidt, S., Grabeel, J., Bankamp, B. (2021). Pilot Investigation of SARS-CoV-2 Secondary Transmission in Kindergarten Through Grade 12 Schools Implementing Mitigation Strategies — St. Louis County and City of Springfield, Missouri, December 2020. MMWR. Morbidity and Mortality Weekly Report, 70(12), 449–455. https://doi.org/10.15585/mmwr.mm7012e4.

Dibner, K. A., Christakis, D. A., & Schweingruber, H. A. (2020, September 1). Reopening K-12 Schools During the COVID-19 Pandemic. JAMA. https://jamanetwork.com/journals/jama/fullarticle/2769036.

Dimitri A. Christakis, M. D. (2020, October 1). School Reopening-The Pandemic Issue That Is Not Getting Its Due. JAMA Pediatrics. https://jamanetwork.com/journals/jamapediatrics/fullarticle/2766113.

Gold, J. A., Gettings, J. R., Kimball, A., Franklin, R., Rivera, G., Morris, E., Scott, C., Marcet, P. L., Hast, M., Swanson, M., McCloud, J., Mehari, L., Thomas, E. S., Kirking, H. L., Tate, J. E., Memark, J., Drenzek, C., Vallabhaneni, S., Almendares, O., … Weng, M. K. (2021). Clusters of SARS-CoV-2 Infection Among Elementary School Educators and Students in One School District — Georgia, December 2020–January 2021. MMWR. Morbidity and Mortality Weekly Report, 70(8), 289–292. https://doi.org/10.15585/mmwr.mm7008e4.

Hershow, R. B., Wu, K., Lewis, N. M., Milne, A. T., Currie, D., Smith, A. R., Lloyd, S., Orleans, B., Young, E. L., Freeman, B., Schwartz, N., Bryant, B., Espinosa, C., Nakazawa, Y., Garza, E., Almendares, O., Abara, W. E., Ehlman, D. C., Waters, K., Hill, M., Chu, V. (2021, April 28). Low SARS-CoV-2 Transmission in Elementary Schools - Salt Lake County, Utah, December 3, 2020–January 31, 2021. Centers for Disease Control and Prevention. https://www.cdc.gov/mmwr/volumes/70/wr/mm7012e3.htm?s_cid=mm7012e3_w.

Krishnaratne, S., Pfadenhauer, L. M., Coenen, M., Geffert, K., Jung-Sievers, C., Klinger, C., Kratzer, S., Littlecott, H., Movsisyan, A., Rabe, J. E., Rehfuess, E., Sell, K., Strahwald, B., Stratil, J. M., Voss, S., Wabnitz, K., & Burns, J. (2020). Measures implemented in the school setting to contain the COVID-19 pandemic: a rapid scoping review. Cochrane Database of Systematic Reviews. https://doi.org/10.1002/14651858.cd013812

Ladhani, S. N., Baawuah, F., Beckmann, J., Okike, I. O., Ahmad, S., Garstang, J., Brent, A. J., Brent, B., Walker, J., Andrews, N., Ireland, G., Aiano, F., Amin-Chowdhury, Z., Letley, L., Flood, J., Jones, S., Borrow, R., Linley, E., Zambon, M., Poh, J., … Ramsay, M. E. (2021). SARS-CoV-2 infection and transmission in primary schools in England in June-December, 2020 (sKIDs): an active, prospective surveillance study. The Lancet. Child & adolescent health, 5(6), 417–427. https://doi.org/10.1016/S2352-4642(21)00061-4

Lanier, W. A., Babitz, K. D., Collingwood, A., Graul, M. F., Dickson, S., Cunningham, L., Dunn, A. C., MacKellar, D., & Hersh, A. L. (2021). COVID-19 Testing to Sustain In-Person Instruction and Extracurricular Activities in High Schools — Utah, November 2020–March 2021. MMWR. Morbidity and Mortality Weekly Report, 70(21), 785–791. https://doi.org/10.15585/mmwr.mm7021e2

Lewis, N. M., Hershow, R. B., Chu, V. T., Wu, K., Milne, A. T., LaCross, N., Hill, M., Risk, I., Hersh, A. L., Kirking, H. L., Tate, J. E., Vallabhaneni, S., & Dunn, A. C. (2021). Factors Associated with Participation in Elementary School–Based SARS-CoV-2 Testing — Salt Lake County, Utah, December 2020–January 2021. MMWR. Morbidity and Mortality Weekly Report, 70(15), 557–559. https://doi.org/10.15585/mmwr.mm7015e1

McGee, R.S., Homburger, J.R., Williams, H.E., Bergstrom, C.T., & Zhou, A.Y. (2021). Model-driven mitigation measures for reopening schools during the COVID-19 pandemic. Proc Natl Acad Sci U S A. 2021 Sep 28;118(39):e2108909118. doi: 10.1073/pnas.2108909118. PMID: 34518375.

Prem, K., Liu, Y., Russell, T. W., Kucharski, A. J., Eggo, R. M., Davies, N., Jit, M., Klepac, P., Flasche, S., Clifford, S., Pearson, C. A., Munday, J. D., Abbott, S., Gibbs, H., Rosello, A., Quilty, B. J., Jombart, T., Sun, F., Diamond, C., … Hellewell, J. (2020). The effect of control strategies to reduce social mixing on outcomes of the COVID-19 epidemic in Wuhan, China: a modelling study. The Lancet Public Health, 5(5). https://doi.org/10.1016/s2468-2667(20)30073-6

Reitsma, M. B., Claypool, A. L., Vargo, J., Shete, P. B., McCorvie, R., Wheeler, W. H., Rocha, D. A., Myers, J. F., Murray, E. L., Bregman, B., Dominguez, D. M., Nguyen, A. D., Porse, C., Fritz, C. L., Jain, S., Watt, J. P., Salomon, J. A., & Goldhaber-Fiebert, J. D. (2021). Racial/Ethnic Disparities In COVID-19 Exposure Risk, Testing, And Cases At The Subcounty Level In California. Health Affairs, 40(6), 870–878. https://doi.org/10.1377/hlthaff.2021.00098

Rogers, R. W., & Prentice-Dunn, S. (1997). Protection motivation theory. In D. S. Gochman (Ed.), Handbook of health behavior research 1: Personal and social determinants (pp. 113–132). Plenum Press.

